# Mathematical Modelling of Oxygenation Dynamics using High-Resolution Perfusion Data – Part 1: Statistical Framework

**DOI:** 10.1101/2024.03.18.24304520

**Authors:** Mansour T. A. Sharabiani, Alireza S. Mahani, Richard W. Issitt, Yadav Srinivasan, Alex Bottle, Serban Stoica

## Abstract

**Background:** Balancing oxygen supply and demand during cardiopulmonary bypass (CPB) is crucial to minimise adverse outcomes. This is managed by adjusting oxygen delivery components – cardiac index (CI), haemoglobin concentration (Hb), arterial oxygen saturation (SaO_2_) – and metabolic demand through temperature (Temp) changes. The oxygen extraction ratio (OER) responds to these adjustments, affecting oxygen consumption, but this response is not well understood. We aimed to develop a mathematical model to capture OER dynamics during CPB and quantify oxygen demand’s dependence on temperature.

**Methods:** We developed GARIX, a time-series model predicting minute-by-minute OER changes during CPB, incorporating exogenous variables (CI, Hb, SaO_2_, Temp) and an equilibrium term representing the difference between oxygen consumption and temperature-dependent oxygen demand, modelled linearly per the van’t Hoff specification (constant Q_10_). The model was trained on data from 343 CPB operations (20,000 minutes) in 334 paediatric patients at a UK centre (2019–2021). We used variable importance analysis and simulations to study the model’s properties.

**Results:** The model shows OER adapts to align oxygen consumption with demand. The adaptive response has a rapid phase (<10 minutes) and a slower phase extending up to several hours. Equilibrium analysis yields Q_10_ = 2.25, indicating oxygen demand doubles with every 8.5°C increase in temperature during CPB.

**Conclusions:** Our model provides a physiologically plausible framework for explaining OER changes during CPB, capturing dynamic adjustments and steady-state oxygen consumption. These findings highlight the value of mathematical modelling in estimating key oxygenation parameters like Q_10_, given limitations on clinical experimentation.

## Introduction

During cardiopulmonary bypass (CPB), the heart-lung machine temporarily assumes the perfusion functions of maintaining oxygenation and circulation. Balancing oxygen supply and demand is critical in goal-directed perfusion (GDP) during CPB. Oxygen supply is managed through cardiac index (CI), haemoglobin concentration (Hb), arterial oxygen saturation of haemoglobin (SaO_2_) and partial pressure of oxygen in arterial blood (PaO_2_), while oxygen demand is dictated by the metabolic rate which is primarily a function of body temperature (Temp) during CPB. Effective GDP hinges on two key questions: (1) What is the body’s oxygen demand at any given time? (2) How does oxygen extraction respond to changes in oxygen delivery and demand?

While CI, Hb, SaO_2_, PaO_2_, and Temp are controlled by the perfusionist, the oxygen extracted by the body is endogenous and cannot be directly manipulated. The assumption of ‘supply-independent consumption’^1^ – that the body consumes only what it needs regardless of supply – may not hold during CPB due to frequent abrupt changes. For example, pump flow may be reduced during surgical manoeuvres, or Hb levels may change due to haemodilution or transfusions. It is unreasonable to presume the body instantaneously adapts to such changes and maintains perfect oxygen balance during CPB.

Existing models that relate Temp to oxygen demand, such as the van’t Hoff and Arrhenius equations, rely on limited human data and are unlikely to be updated due to ethical constraints. Notably, the ‘textbook’ model^2^ is based on only two temperature points at 20°C in humans^3^ and 37°C in animals^4–6^.

High-resolution observational data collected during CPB present new opportunities to study oxygenation dynamics but also pose challenges. Traditional studies often assume the body is in equilibrium – a condition not met during CPB. Therefore, sophisticated statistical techniques are necessary to analyse such data accurately, accounting for system dynamics while attempting to infer the body’s latent oxygen demand in equilibrium.

In this study, we introduce a global autoregressive integrated time-series model with exogenous variables and an equilibrium term (GARIX) to predict minute-by-minute changes in oxygen extraction ratio (OER), which is the fraction of arterial oxygen extracted by the body. The model accounts for past changes in the system, its current state, and the intended perfusionist actions. To assess the model’s physiological plausibility, we examine its coefficients, perform variable importance analysis, and conduct simulations of system behaviour under various scenarios. For a list of nonstandard abbreviations and acronyms used in this manuscript, see Supplementary Material A.

### Patients and Methods

#### Study Design and Data Collection

We conducted a retrospective analysis of high-resolution intraoperative data from paediatric patients undergoing cardiopulmonary bypass (CPB) operations at a single centre in the UK between 2019 and 2021. Institutional approval was obtained from the Great Ormond Street Hospital for conducting this study (audit number 3045). Clinical data were collated in a research platform within the hospital’s governance structure and de-identified before analysis. Due to the retrospective nature of the study, the NHS Health Research Authority London—Bloomsbury Research Ethics Committee (REC reference: 17/LO/0008) waived the need for obtaining informed consent. All methods were performed in accordance with the relevant guidelines and regulations.

Continuous minute-by-minute measurements were collected, including CI, Hb, SaO_2_, SvO_2_, and Temp. To ensure physiological insights reflected non-pathological responses, we excluded patients who developed postoperative acute kidney injury (AKI), resulting in a training dataset of 334 patients undergoing 343 CPBs with a total operation time of 19,687 minutes. Detailed data collection and preparation methods are provided in Supplementary Material B.

### Mathematical Modelling of Oxygen Extraction Dynamics

#### Conceptual Framework of the GARIX Model

To predict minute-by-minute changes in the oxygen extraction ratio (OER) during CPB, we developed a novel, physiology-inspired time-series model called the Global Autoregressive Integrated model with Exogenous variables and an Equilibrium term (GARIX). The model captures the dynamic interplay between oxygen delivery, oxygen demand, and the body’s adaptive responses by integrating three key components:

- **History Term Group (HTG)**: Accounts for the influence of past changes in exogenous variables and OER on current OER dynamics.
- **Equilibrium Term Group (ETG)**: Represents the body’s effort to balance oxygen consumption with temperature-dependent oxygen demand.
- **Future Term Group (FTG)**: Reflects the impact of intended immediate changes in exogenous variables due to clinical interventions.

By incorporating these components, the GARIX model provides a comprehensive framework for understanding how OER adapts over time in response to both physiological processes and perfusionist actions.

#### Model Specification

The model predicts changes in the logit-transformed OER from time t to t+1 minute as:

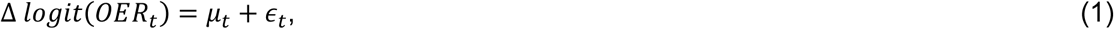

where:

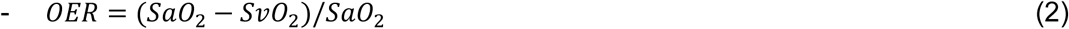
– Δ*logit*(*OER*_*t*_) = *logit*(*OER*_*t*+1_) − *logit*(*OER*_*t*_),
– μ_*t*_ is the expected change in OER,
– ε_*t*_ is a random error term assumed to be normally distributed with zero mean and constant variance,
– The logit function is defined as *logit*(*x*) = ln(^*x*/1 − x^).

#### First Perspective: Decomposition into History, Equilibrium, and Future Term Groups

In the first perspective, the expected change μ_*t*_ is composed of three term groups:

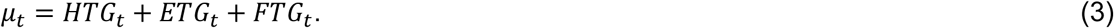

#### History Term Group (HTG)

The HTG captures the cumulative effect of past changes over N previous minutes:

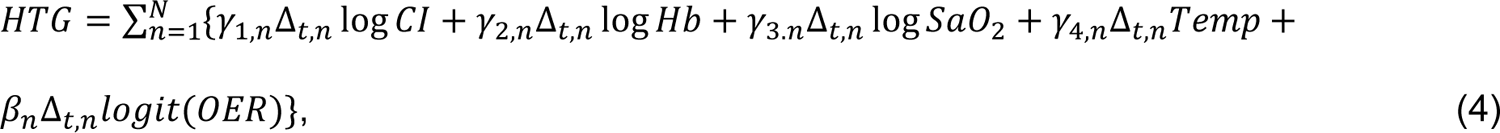

where:

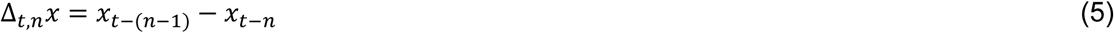

represents the change in variable *x* from *t* − *n* to *t* − (*n* − 1) and γ_*i,n*_ and β_*n*_ are model coefficients for each variable and lag *n*.

#### Equilibrium Term Group (ETG)

The ETG embodies the body’s drive to align oxygen consumption (VO_2_i) with temperature-dependent oxygen demand (tVO_2_i):

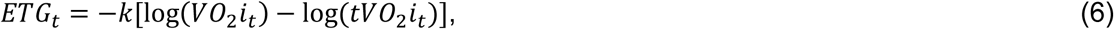

where,

– k is a positive coefficient representing the strength of the equilibrium force,

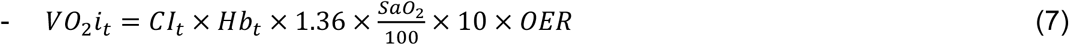
– is the indexed oxygen consumption at time t,
– *tVO*_2_*i*_*t*_ is the target (indexed) oxygen consumption, modelled following van’t Hoff specification:

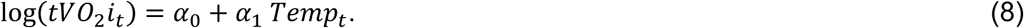

In steady-state conditions, ETG = 0, or consumption equals demand. This means:

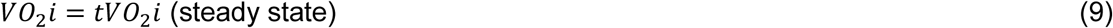

If CI, Hb, SaO_2_ and Temp are given, and α_0_, α_1_ are estimated from data, we can combine Equations 7-9 to solve for OER.

#### Future Term Group (FTG)

The FTG accounts for immediate intended changes due to clinical interventions from time t to t+1:

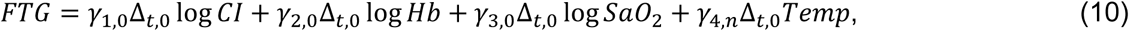

with Δ_*t*,0_*x* = *x*_*t*+1_ − *x*_*t*_ representing the intended change in variable *x* at time t.

Note: The term involving *logit*(*OER*) is absent because it represents the outcome variable we aim to predict.

#### Second Perspective: Decomposition into Autoregressive, Exogenous, and Equilibrium Term Groups

Alternatively, the expected change μ_*t*_ can be decomposed into:

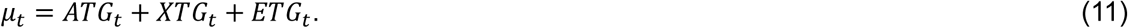

#### Autoregressive Term Group (ATG)

The ATG captures the influence of past changes in OER on its future trajectory:

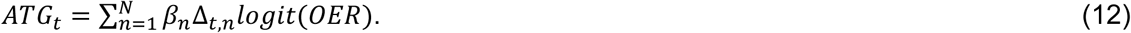

Interpretation: Negative coefficients β_*n*_ imply a self-correcting mechanism in OER dynamics.

#### Exogenous Term Group (XTG)

The XTG represents the effects of changes in exogenous variables (CI, Hb, SaO_2_, Temp):

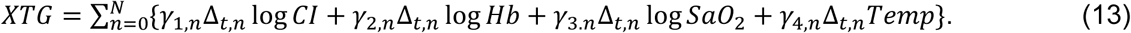

Note: Includes both historical (n > 0) and immediate (n=0) changes. The ETG is the same as the first perspective.

#### Relationship Between the Two Perspectives

– The Equilibrium Term Group (ETG) is common to both decompositions.
– The History Term Group (HTG) and Future Term Group (FTG) can be rearranged to form the Autoregressive Term Group (ATG) and Exogenous Term Group (XTG):

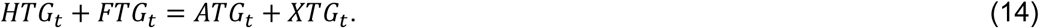
– This dual perspective allows us to interpret the model both in terms of physiological mechanisms (history, equilibrium, future actions) and statistical time-series components (autoregressive, exogenous variables).

#### Parameter Estimation

The parameters γ_*i,n*_, β_*n*_, *k*, α_0_, α_1_ were estimated using linear regression techniques on the training dataset. Detailed derivations, including transformations and interpretations of parameters, are provided in Supplementary Material C.

#### Model Training and Selection

We estimated the model coefficients using linear regression techniques on the training dataset. The history length N was treated as a hyperparameter, selected by maximizing cross-validated R-squared performance. We found that a history length of N=7 minutes provided a balance between model complexity and predictive accuracy, leading to the GARIX(7) model.

#### System Simulations

We performed deterministic and stochastic simulations to explore the model’s behaviour under various scenarios. Detailed methodologies and results are available in Supplementary Material G.

#### Model Evaluation and Diagnostics

Model performance was assessed using cross-validation, and variable importance was analysed using permutation-based methods (details in Supplementary Material E).

## Results

We analysed data from 343 operations in patients without AKI, focusing on their intraoperative oxygenation dynamics. Supplementary Material I provides the details of patients and outcomes.

### Adaptive OER Response

Figure 1 (panels A–E) displays the estimated means and 95% confidence intervals for the coefficients of all state variables (OER, CI, Hb, SaO_2_, Temp) in a GARIX(20) model, plotted against term lag *n*, defined in Equation 5. Coefficient magnitudes decrease with increasing lag and become – mostly – statistically insignificant after 10 minutes. Panel F shows that the model’s out-of-sample (OOS) performance does not meaningfully improve beyond 5 terms, consistent with the coefficient trends. Therefore, we chose GARIX(7) for subsequent analyses. (Note that this does not imply the system reaches a new equilibrium within 10 minutes after a perturbation, as discussed later.)

**Figure 1:**
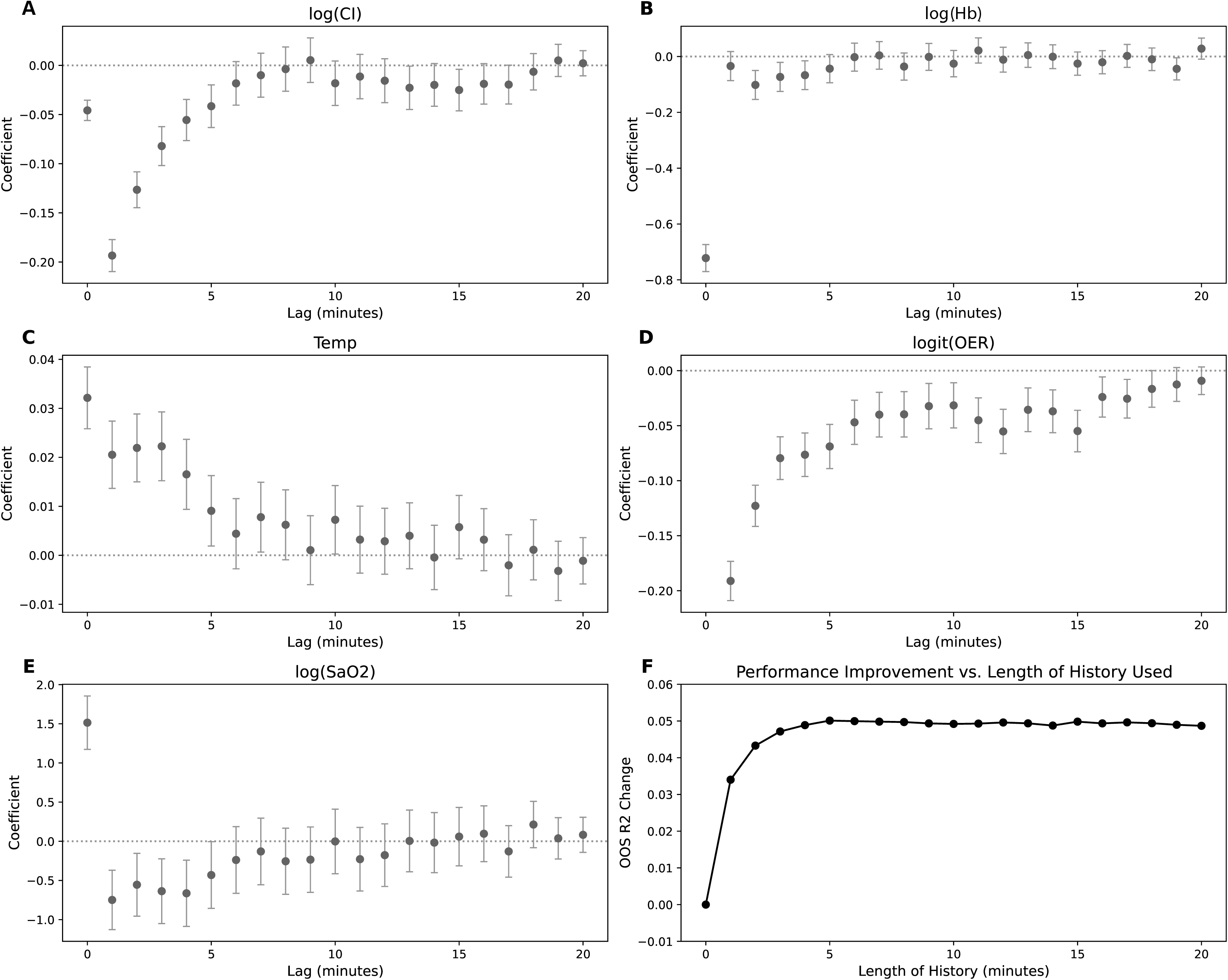
Adaptive OER Response Length of history vs. model coefficients and performance. Panels A-E show the coefficients of different state variables vs. the lag order (index ‘n’ in Equation 4). Error bars represent 95% confidence intervals for each coefficient. Panel F shows change in OOS R-squared – compared to R-squared for N = 0 – vs. N (or maximum of ‘n’ in Equations 4) in GARIX(N).

The signs of the coefficients for the state variables are consistent with an adaptive *OER* response that: (1) maintains steady oxygen consumption when temperature remains constant; (2) increases or decreases oxygen consumption when temperature increases or decreases, respectively; and (3) stabilizes random fluctuations in oxygen consumption.

#### CI and Hb

If CI increases while Hb, SaO_2_ and Temp remain unchanged, oxygen delivery rises while oxygen demand remains the same. Assuming oxygen supply and demand were balanced before the CI increase, the body must reduce OER to lower consumption and retain the balance. This necessitates a negative coefficient for CI change terms, which we observe in Figure 1A, indicating a physiologically adaptive OER response. An identical argument applies to Hb, confirmed by the negative coefficients in Figure 1B.

#### Temp

An increase in Temp elevates oxygen demand, requiring an increase in OER to match consumption with the higher demand. Thus, we expect positive coefficients for Temp-related terms, as seen in Figure 1C.

#### OER

If OER increases without a change in Temp, oxygen consumption would exceed demand. To restore balance, delivery must decrease. Therefore, the autoregressive coefficients should be negative, as shown in Figure 1D.

#### SaO_2_

SaO_2_ is unique for two reasons. First, it exhibits a very narrow dynamic range in our data (interquartile range 98%–99%), partly due to rounding errors. Second, SaO_2_ directly influences OER given its definition. The significant positive coefficient at lag 0 (Figure 1E) reflects this direct effect: an increase in SaO_2_, with no immediate change in SvO_2_, increases OER given the definition in Equation 2. The marginally significant negative coefficients at lags 1 and 3 suggest an adaptive response like that for CI and Hb, where OER decreases to counteract the rise in SaO_2_ and maintain steady consumption.

#### Timescale of Adaptations

Figure 2A shows the contributions of different TGs to the predicted changes in *logit*(*OER*) over time, following a simulated 25% decrease in CI at time *t* = 0. Simulations for panels A and B were conducted in deterministic mode. The initial rapid adaptation is primarily driven by the XTG, while the subsequent slower adaptation is governed by the counteracting effects of the ETG and ATG. The ATG partially offsets the ETG, resulting in a slower overall adaptation rate. The resulting OER trajectory is illustrated in Figure 2B (“original” series).

**Figure 2:**
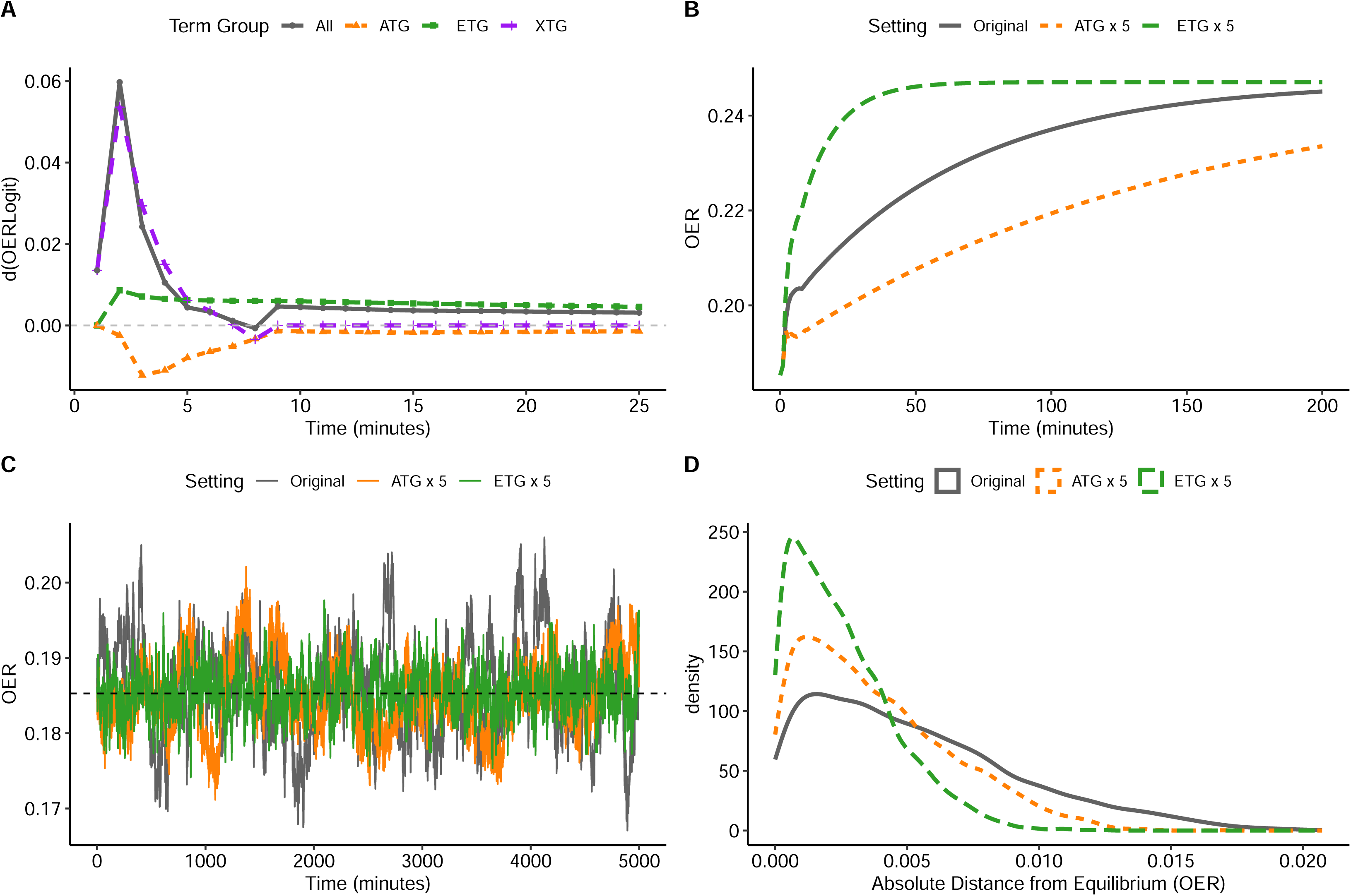
Timescale of OER Adaptation Timescale of OER adaptation. A: Components of change – by term group – in logit of OER over time, after a 25% decrease in CI at t=0. Prior to this step change, the system is in equilibrium at t = 0, with state variables being log *CI*= 1.00, log *Hb*= 2.31, log *SaO*_2_= 4.60, *T* = 33.1*C*. B: *OER* trajectory (‘original’ series, grey), corresponding to the same experiment as in panel A. The y values are the inverse logit of the cumulative sum of the ‘total’ time series in the left panel. The ‘ATG x 5’ series (orange) represents OER trajectory for a hypothetical system where the coefficients of all autoregressive terms are multiplied by 5. The ‘ETG x 5’ series (green) represent a hypothetical system with the equilibrium coefficients multiplied by 5. C: OER trajectory using stochastic simulations and same initial values for state variables prior to *CI* change at t=0. D: Histogram of absolute distance from equilibrium OER for the same data as in panel C. For panels C and D, noise level used in simulations was set to 1/10^th^ of the value estimated from the data.

To further demonstrate the counteracting effects of the ATG and ETG during the transition to a new equilibrium, panel B also includes simulations where the coefficients of the ATG (“ATG × 5”) or the ETG (“ETG × 5”) are amplified. Amplifying the ATG terms slows down the transition, whereas amplifying the ETG terms accelerates it.

Conversely, when the system is already at equilibrium, the ATG and ETG forces align, both acting to dampen stochastic fluctuations in OER around the equilibrium point. Panel C presents a sample trajectory – using stochastic simulation mode – for the same three scenarios as in panel B. The reduction in mean absolute deviation from the equilibrium OER, after amplifying the ATG or ETG forces, is more clearly depicted in the kernel density plots of panel D.

#### Oxygen Demand vs. Temperature

Consider a state of perfect equilibrium where all variables, including OER, remain constant over time. In this situation, the expected change in *logit*(*OER*), denoted as μ_*t*_ in Equation 11, must be zero for all time points *t*. Since the ATG and XTG consist solely of change terms, they become zero in equilibrium. For the ETG to also be zero, *log*(*VO*_2_*i*) must equal *log*(*tVO*_2_*i*). This implies that, at equilibrium, oxygen consumption matches oxygen demand.

According to the van’t Hoff model we have adopted, the logarithm of oxygen demand is a linear function of temperature (Equation 8). Therefore, in fitting GARIX to the data, the parameters of the van’t Hoff equation are also estimated, quantifying how the body’s latent oxygen demand during CPB depends on temperature.

Using Equations C.16a-c (Supplementary Material C) and Monte Carlo simulations, we estimated median and confidence interval values for the parameters *k*, α_0_, and α_1_. The 95% confidence interval (CI) for *k* is [0.026 – 0.037] (median: 0.031), which is positive as expected, justifying its interpretation as a (return to) equilibrium force. Applying Equation C.14, we estimated the 95% CI for *Q*_10_ to be [1.83 – 2.73] (median: 2.25). For comparison, the median estimate of *Q*_10_ from a naïve (static) model fitted to the same data (see Supplementary Material H) is significantly higher at 3.47, with a 95% CI of [3.42 – 3.52]. For comparison, using a combination of human and animal experiments, Fox et al (1982) estimated Q_10_ to be 2.4, which is quite close to our estimate of 2.25, using GARIX.

The large difference between the Q_10_ estimated by GARIX and the value resulting from a naïve fit to the oxygen consumption data reflects systemic gaps between the observed oxygen consumption and the latent oxygen demand. Figure 3 illustrates this point, suggesting under-oxygenation at low temperatures (< 32C) and over-oxygenation at high temperatures (> 32C). Given that most temperature recordings are above 32C (69%), we observe an overall over-oxygenation in our data by ∼8%.

**Figure 3:**
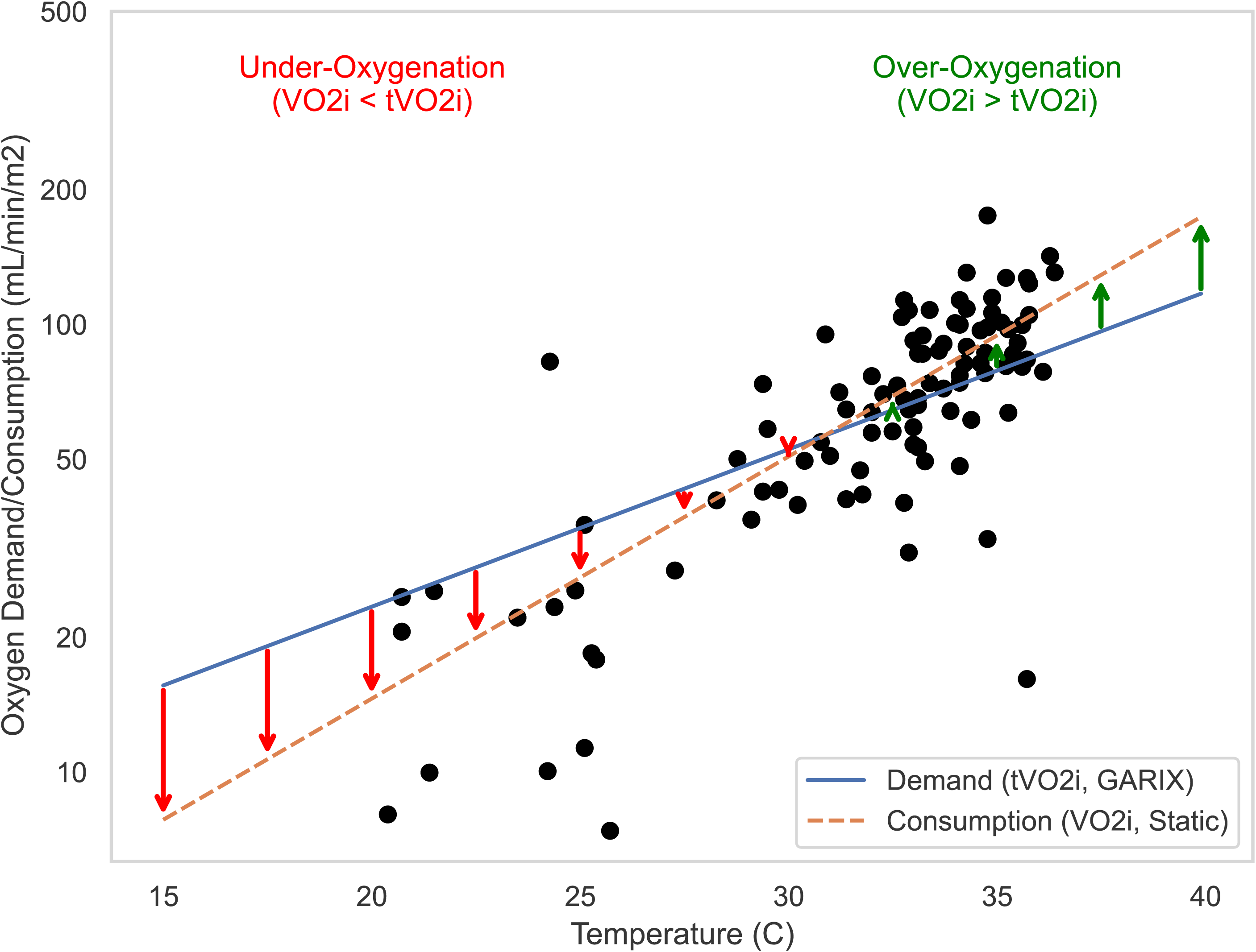
Systemic Oxygenation Gap vs. Temperature Systemic oxygenation gap vs. temperature. Blue line is the GARIX(7)-estimated oxygen ‘demand’ vs temperature. Grey dots are a random 100 pairs of temperature and oxygen consumption from the data. Orange line is a log-linear fit to the data (see Naïve model in Supplementary Material H) and represents the average oxygen ‘consumption’ at each temperature. The arrows indicate the magnitude and direction of systemic oxygenation gap (between consumption and demand) at different temperatures.

#### Variable Importance Analysis

Permutation-based variable importance (PVI) for history, action (or future) and equilibrium (or present) term groups are 6.7% +/- 0.5%, 6.0% +/- 0.5% and 0.78% +/- 0.07%, respectively. (See Supplementary Material F for details.) The equilibrium term makes the smallest, yet statistically significant, contribution to predictive power, highlighting the importance of our large training data for GARIX. About a half of total predictive power is due to the historical changes, reflecting significant system memory and the importance of multi-horizon planning for controlling OER during CPB.

#### Comment

This study introduces the GARIX model, a novel data-driven analytical framework for predicting changes in the OER in response to clinical interventions during CPB. Our analysis indicates that OER adapts slowly to maintain balance between oxygen supply and demand. Notably, the inclusion of an equilibrium force GARIX allowed us to estimate how oxygen demand during CPB depends on body temperature, quantified by the *Q*_10_ parameter.

#### GARIX as a Grey-Box Model

In developing GARIX, we enhanced the traditional autoregressive integrated time-series model with exogenous variables (ARIX) in two significant ways. First, GARIX is a ‘global’ model combining data from different patients and procedures, providing sufficient power to estimate ETG parameters despite its relatively low predictive power. However, this produces a single set of coefficients, not accounting for differences in patient characteristics like age or operational variations. Future work will address this by incorporating these variables or estimating models for subgroups.

Second, GARIX includes the equilibrium term group (ETG), analogous to cointegration and error correction in econometrics^9^. The ETG introduces an adaptive mechanism ensuring oxygen consumption matches metabolic needs. Without it, GARIX would accept any constant combination of state variables as steady-state. By assuming oxygen demand depends on temperature via the van’t Hoff model, we incorporate domain knowledge into the statistical framework, making GARIX a ‘grey box’ model.

Our large dataset was crucial for this modelling approach. GARIX(7) contains 40 coefficients and is estimated using nearly 20,000 data points. Most explanatory power comes from dynamic terms, which is why the confidence intervals for *Q*_10_ are significantly wider than those from a naïve static model. A smaller dataset might not have allowed us to estimate *Q*_10_ with statistical significance.

As this is, to our knowledge, the first model predicting OER changes during CPB, we prioritized ‘explainability’. The linear specification of GARIX enabled us to examine and interpret the model coefficients, ensuring their physiological plausibility. Such restrictions will be relaxed in future work.

#### OER Adaptation and Its Timescales

Our simulations indicated that the oxygen extraction ratio (OER) adapts to changes in oxygen delivery over distinct timescales: An initial rapid adaptation primarily driven by the XTG, followed by a slower adaptation involving counteracting effects between the ETG and the ATG.

One potential physiological mechanism for the fast adaptation phase, occurring within minutes, is microvascular regulation. Rapid vasodilation or vasoconstriction of arterioles and capillaries may adjust blood flow to match regional metabolic demands, helping to mitigate immediate imbalances in oxygen delivery^10^. This swift response could reflect the body’s initial effort to maintain homeostasis during abrupt changes in perfusion parameters.

The slower adaptation phase, extending over several hours, might be associated with changes in gene expression. Cells could alter the expression of genes involved in metabolic pathways and oxygen utilization, leading to longer-term adjustments in oxygen consumption^11^.

The resistance effect of the ATG during the transition to a new equilibrium might physiologically serve to prevent abrupt changes in OER that could be harmful. By slowing down the adaptation, the body may be avoiding rapid shifts that exceed its capacity to compensate safely.

### Model Assumptions and Limitations

#### Dissolved Oxygen

Our model does not account for the contribution of dissolved oxygen to oxygen extraction due to the lack of data on PvO_2_. However, dissolved oxygen constitutes, on average, only 1.6% of the total arterial oxygen content in our data. We therefore consider our analysis directionally correct. Future work will incorporate this component to improve model precision.

#### Exogeneity vs. Endogeneity

We assumed that Temp, CI, Hb, SaO_2_ are exogenous variables, which is an idealisation. These variables at time *t* + 1 cannot be set with certainty at time *t*; they are controlled indirectly, blurring the line between exogeneity and endogeneity. For example, SaO_2_ is influenced by arterial oxygen pressure (PaO_2_). A basic dynamic model (results not shown) confirmed that PaO_2_ and Temp can predict changes in SaO_2_ accurately. Future models should better reflect the true controllability of these variables.

#### Risk of Extrapolation

Apart from using a logit transformation to constrain OER between 0 and 1, we imposed no other constraints. The nonlinearity of the logit function naturally slows changes in OER near extreme values, adding physiological plausibility to our simulations.

Perfusionists strive to maintain OER within tighter ranges; in our data, OER remains below 31% for 90% of the time. Therefore, caution is needed when interpreting simulation results where OER approaches or exceeds the observed range in the training data.

#### Linearity

We chose a linear model to ensure interpretability and explainability, given the physiological complexity of OER dynamics. While more sophisticated, e.g., machine learning, approaches could capture nonlinear relationships and potentially improve predictive performance, our priority was to establish a transparent baseline model. Linear models allow direct interpretation of how exogenous variables – CI, Hb, SaO_2_ and Temp – affect OER changes, which is challenging with complex, black-box models.

#### Homoscedasticity

Our model assumes homoscedasticity – that the variance of residuals is constant across all levels of predicted OER. However, residual analysis (Supplementary Material H) suggests the presence of heteroscedasticity. Our model may thus not fully capture the heterogeneity of physiological noise, especially under stress or abrupt changes in exogenous factors like CI or Hb. Future iterations could address heteroscedasticity using generalized least squares or more flexible models, enhancing accuracy and robustness by accounting for variability across different system states.

## Conclusions

In this study, we used high-resolution paediatric CPB data to develop a mathematical model predicting minute-by-minute changes in oxygen extraction in response to perfusionist interventions during CPB. The model coefficients indicate an adaptive response aimed at balancing oxygen supply and demand; however, the timescale of these adaptations leaves ample room for short-term imbalances. By analysing the model’s equilibrium characteristics, we estimated the dependence of the body’s oxygen demand on temperature during CPB, including the *Q*_10_ parameter. Building on the analytical framework laid in this paper, future work will delve deeper into the physiological implications of GARIX and explore its clinical implications.

## Declaration of Generative AI and AI-assisted technologies in the writing process

During the preparation of this work the author(s) used ChatGPT in order to edit and reorganise the manuscript. After using this tool/service, the author(s) reviewed and edited the content as needed and take(s) full responsibility for the content of the publication.

## Data Availability

All data produced in the present study are available upon reasonable request to the authors

