## Supplementary Material for "Mathematical Modelling of Oxygenation Dynamics using High-Resolution Perfusion Data – Part 1: Statistical Framework"

**Brief Title**: Paediatric CPB Oxygenation Dynamics

**Authors**: Mansour T. A. Sharabiani, PhD[a†], Alireza S. Mahani, PhD[b†], Richard W. Issitt, DClinP[c,d,e†], Yadav Srinivasan, MD[f], Alex Bottle, PhD[a], Serban Stoica, FRCS MD[g].

**Affiliations**:

a. School of Public Health, Imperial College, London, United Kingdom

b. Statman Solution Ltd., London, United Kingdom

c. Centre for Heart Failure, Transplantation and Extracorporeal Support, UCL Institute for Cardiovascular Science, UCL, London, United Kingdom

d. Perfusion Department, Great Ormond Street Hospital for Children, London, United Kingdom

e. Data Research, Innovation and Virtual Environments Unit, Great Ormond Street Hospital for Children, London, United Kingdom

f. Cardiac Surgery Department, Great Ormond Street Hospital for Children, London, United Kingdom

g. Cardiac Surgery Department, Bristol Royal Children’s Hospital, Bristol, United Kingdom

† These authors contributed equally to this work.

The authors have no conflicts of interest to disclose.

**Corresponding Author**:

Dr Mansour T A Sharabiani

School of Public Health, Imperial College London

London, United Kingdom, W12 0BZ

**Corresponding Co-Author**:

Dr Richard Issitt

Centre for Heart Failure, Transplantation and Extracorporeal Support, Research Department of Children's Cardiovascular Disease, UCL Institute of Cardiovascular Science, London, United Kingdom.

### Supplementary Materials

#### A. Nonstandard Abbreviations and Acronyms

- AKI: acute kidney injury
- ATG: autoregressive term group
- BSA: body surface area ($m^2$)
- CI: cardiac index (blood flow indexed to BSA; $L/min/m^2$)
- CPB: cardiopulmonary bypass
- CV: cross-validation
- $DO_2$: oxygen delivery (mL/min)
- $DO_2i$: oxygen delivery, indexed to BSA ($mL/min/m^2$)
- ETG: equilibrium term group
- FTG: Future Term Group

- GARIX: A global autoregressive integrated (time-series) model with exogenous variables, plus a disequilibrium term group
- GDP: goal-directed perfusion
- Hb: haemoglobin content of blood (g/dL)
- Hct: haematocrit
- HTG: History Term Group
- i.i.d: independent and identically distributed
- KDIGO: Kidney Disease: Improving Global Outcomes
- OER: oxygen extraction ratio
- OOS: out-of-sample
- $PaO_2$: partial pressure of oxygen in arterial blood (kPa)
- $PvO_2$: partial pressure of oxygen in mixed venous blood (kPa)
- PRAiS2: partial risk adjustment in surgery model
- PTG: Present Term Group
- PVI: permutation-based variable importance
- Q10: multiplicative increase in the resting metabolic rate for every 10℃ increase in body temperature
- RMR: resting metabolic rate
- $SaO_2$: oxygen saturation of arterial blood (%)
- $SvO_2$: oxygen saturation of mixed venous blood (%)
- TG: term group
- $tVO_2i$: target (indexed) oxygen consumption (indexed oxygen demand; $mL/min/m^2$)
- VI: variable importance
- $VO_2$: oxygen consumption (mL/min)
- $VO_2i$: oxygen consumption, indexed to BSA ($mL/min/m^2$)
- XTG: Exogenous Term Group

### B. Study Design and Data Preparation

#### Data Collection

Intraoperative data from the heart-lung machine and anaesthetic monitors were captured every minute throughout surgery. Continuous measurements included:

- **Cardiac Index (CI)**: Blood flow indexed to body surface area (BSA), measured in $L/min/m^2$.
- **Haemoglobin Concentration (Hb)**: Measured in g/dL.
- **Arterial Oxygen Saturation ($SaO_2$)**: Percentage of oxygen-saturated haemoglobin in arterial blood.
- **Venous Oxygen Saturation ($SvO_2$)**: Percentage of oxygen-saturated haemoglobin in mixed venous blood.
- **Body Temperature (Temp)**: Measured in degrees Celsius (°C).

Intraoperative blood biochemistry was continuously monitored using an optical fluorescence and reflectance system (CDI550, Terumo, Leuven, Belgium), calibrated every 20 minutes with a cassette-based blood gas analyser (ABL90 Flex Plus, Radiometer, Copenhagen, Denmark).

#### Data Preparation

Data preparation and analysis for this research was conducted using a combination of R and Python programming languages and their various packages.

#### Data Segmentation and Validation

- Each operation was segmented into one or more continuous sessions of minute-by-minute data.
- A validity flag was assigned to each data point based on the following criteria:
    - Haematocrit (Hct) between 0 and 0.6.
    - $SvO_2$ between 0% and 100%.

- $SvO_2$ less than $SaO_2$.
- Temp greater than 0°C.
- CI greater than 0 L/min/m² (to exclude periods of deep hypothermic cardiac arrest).

#### Handling Missing Data and Outliers

- Data points with missing values or those failing the validity criteria were excluded from analysis.
- Records at the start and end of each operation with CI = 0 (due to data-recording processes) were removed.
- Only valid, contiguous time segments were used for model training.

#### Derived Variables

- **Body Surface Area (BSA)**: Calculated using the Mosteller formula:

$$BSA = \sqrt{\frac{Height \times Weight}{3600}}, \quad \text{(B.1)}$$

where height is in centimetres and weight is in kilograms.

- **Indexed Oxygen Delivery ($DO_2i$)**:

$$DO_2i = CI \times Hb \times 1.36 \times (SaO_2/100) \times 10. \quad \text{(B.2)}$$

- **Indexed Oxygen Consumption ($VO_2i$)**:

$$VO_2i = CI \times Hb \times 1.36 \times \left(\frac{SaO_2}{100}\right) \times 10 \times OER. \quad \text{(B.3)}$$

*Note:* The contribution of dissolved oxygen was ignored due to its minimal impact (approximately 1.6% of total arterial oxygen content).

- **Oxygen Extraction Ratio (OER)**:

$$OER = \frac{VO_2i}{DO_2i} = (SaO_2 - SvO_2)/SaO_2. \quad \text{(B.4)}$$

### Final Dataset

- After data preparation, the final training dataset comprised 19,687 minutes of valid observations from 343 operations on 334 patients.
- This dataset was used to train the GARIX(7) model.
- For GARIX(20), similar steps were followed but due to longer history imposing more stringent conditions the training data was smaller at 15,138 minutes.
- When comparing GARIX models of different N's up to 20 (Figure 1F), the training data constructed for GARIX(20) – with a suitable subset of columns in each case - was used for all models to maximize comparability of results.

### C. Detailed Explanation of GARIX Model

### Overview

The GARIX model is designed to predict minute-by-minute changes in the oxygen extraction ratio (OER) during cardiopulmonary bypass (CPB). It incorporates both physiological understanding and statistical modelling techniques. This supplement provides a detailed mathematical formulation of the model, explaining both perspectives:

- **First Perspective**: Decomposition into History Term Group (HTG), Equilibrium Term Group (ETG), and Future Term Group (FTG).
- **Second Perspective**: Decomposition into Autoregressive Term Group (ATG), Exogenous Term Group (XTG), and Equilibrium Term Group (ETG).

We also include parameter transformations, derivations, and interpretations to provide a comprehensive understanding of the model.

### Notation and Definitions

#### *Variables*

- CI: Cardiac Index (L/min/m²)
- Hb: Haemoglobin concentration (g/dL)
- $SaO_2$: Arterial oxygen saturation (%)
- $SvO_2$: Venous oxygen saturation (%)
- Temp: Body temperature (°C)
- OER: Oxygen extraction ratio (between 0 and 1)
- $VO_2i$: Indexed oxygen consumption (mL/min/m²)
- $tVO_2i$: Target indexed oxygen consumption (or oxygen demand) (mL/min/m²)

#### *Operators*

- log: Natural logarithm
- $logit(x) = \log(\frac{x}{1-x})$
- $\Delta_{t,n}x = x_{t-n+1} - x_{t-n}$: Change operator over lag n

### Model Specification

#### *Predicted Change in OER*

The primary equation predicts the change in the logit-transformed OER:

$$\Delta logit(OER_t) = \mu_t + \epsilon_t, \tag{C.1}$$

where $\epsilon_t$ is a random error term assumed to be normally distributed with zero mean and constant variance ($\epsilon_t \sim N(0, \sigma^2)$).

#### *Dual Perspectives of $\mu_t$*

##### First perspective: Decomposition into HTG, ETG and FTG

$$\mu_t = HTG_t + ETG_t + FTG_t. \tag{C.2}$$

**History Term Group (HTG)**:

$$HTG = \sum_{n=1}^{N}\{\gamma_{1,n}\Delta_{t,n}\log CI + \gamma_{2,n}\Delta_{t,n}\log Hb + \gamma_{3.n}\Delta_{t,n}\log SaO_2 + \gamma_{4,n}\Delta_{t,n}Temp + \beta_n\Delta_{t,n}logit(OER)\}. \quad (C.3)$$

**Equilibrium Term Group (ETG)**:

$$ETG_t = -k[\log(VO_2 i_t) - \log(tVO_2 i_t)]. \quad (C.4)$$

- k: Positive equilibrium coefficient.
- $VO_2 i_t$: Calculated as per Equation (C.11).
- $tVO_2 i$: Defined in Equation (C.13).

**Future Term Group (FTG)**:

$$FTG = \gamma_{1,0}\Delta_{t,0}\log CI + \gamma_{2,0}\Delta_{t,0}\log Hb + \gamma_{3,0}\Delta_{t,0}\log SaO_2 + \gamma_{4,n}\Delta_{t,0}Temp. \quad (C.5)$$

### Second Perspective: Decomposition into ATG, XTG, and ETG

$$\mu_t = ATG_t + XTG_t + ETG_t. \quad (C.6)$$

**Autoregressive Term Group (ATG)**:

$$ATG_t = \sum_{n=1}^{N}\beta_n\Delta_{t,n}logit(OER). \quad (C.7)$$

**Exogenous Term Group (XTG)**:

$$XTG = \sum_{n=0}^{N}\{\gamma_{1,n}\Delta_{t,n}\log CI + \gamma_{2,n}\Delta_{t,n}\log Hb + \gamma_{3.n}\Delta_{t,n}\log SaO_2 + \gamma_{4,n}\Delta_{t,n}Temp\}. \quad (C.8)$$

Equilibrium term group (ETG) is the same as in Equation (C.4).

### Relationship Between Perspectives

- The ETG is common to both perspectives.
- The HTG and FTG from the first perspective can be rearranged to form the ATG and XTG:

$$HTG_t + FTG_t = ATG_t + XTG_t. \tag{C.9}$$

### Detailed Calculations

#### *Indexed Oxygen Consumption and Delivery*

Note: The equations below focus on haemoglobin-bound oxygen and ignore the small contribution of dissolved oxygen to the total oxygen content of blood.

Indexed Oxygen Delivery ($DO_2i$):

$$DO_2i_t = CI_t \times Hb_t \times 1.36 \times \frac{SaO_2}{100} \times 10. \tag{C.10}$$

Indexed Oxygen Consumption ($VO_2i$):

$$VO_2i_t = CI_t \times Hb_t \times 1.36 \times \frac{(SaO_2 - SvO_2)}{100} \times 10. \tag{C.11}$$

Since $OER = (SaO_2 - SvO_2)/SaO_2$, we can re-express the above:

$$VO_2i_t = CI_t \times Hb_t \times 1.36 \times \frac{SaO_2}{100} \times 10 \times OER. \tag{C.12}$$

Target Oxygen Consumption ($tVO_2i$):

Following the van't Hoff specification:

$$\log(tVO_2i) = \alpha_0 + \alpha_1\, Temp. \tag{C.13}$$

Calculation of $Q_{10}$:

$$Q_{10} = \exp\,(10\, \alpha_1). \tag{C.14}$$

#### *Reformulation of ETG for linear regression:*

To enable linear regression, we rewrite the ETG:

$$ETG_t = \eta_0 + \eta_1 \log(VO_2i_t) + \eta_2\, Temp_t. \tag{C.15}$$

where:

- $\eta_0 = -k\alpha_0$,
- $\eta_1 = -k$,
- $\eta_2 = -k\alpha_1$.

Recovering original parameters:

- $k = \eta_1$, (C.16a)
- $\alpha_0 = -\eta_0/\eta_1$, (C.16b)
- $\alpha_1 = -\eta_2/\eta_1$. (C.16c)

Monte Carlo Simulations:

Due to the nonlinear transformations, we used Monte Carlo simulations to estimate confidence intervals for $k, \alpha_0, \alpha_1$. Samples were drawn from the joint normal distribution of $(\eta_0, \eta_1, \eta_2)$ obtained from the regression coefficients.

### Interpretation of Term Groups

#### *History Term Group (HTG)*

Purpose: Captures the delayed physiological responses by accounting for historical changes in both exogenous variables and OER.

Components:

- Exogenous variables: $\gamma_{i,n}\Delta_{t,n}x$ terms represent the effect of past changes in exogenous variables on OER.
- Autoregressive component: $\beta_n\Delta_{t,n}logit(OER)$ accounts for influence of past changes in OER itself.

#### *Future Term Group (FTG)*

Purpose: Reflects the intended immediate changes due to clinical interventions.

Components: Immediate (from t to t+1) changes in exogenous variables.

#### *Autoregressive Term Group (ATG)*

Purpose: Models the self-correcting behaviour of OER over time.

Components: Negative $\beta_n$ coefficients indicate that an increase in OER in the past leads to a decrease in the current OER change, promoting stability.

#### *Exogenous Term Group (XTG)*

Purpose: Represents the cumulative effect of changes in exogenous variables on OER.

Components: Includes both historical and immediate changes, encompassing the entire influence of exogenous factors.

#### *Equilibrium Term Group (ETG)*

Purpose: Acts as a corrective force driving the system toward equilibrium where oxygen consumption matches demand.

Interpretation: Positive k ensures that when $VO_2i > tVO_2i$ (over-oxygenation), the ETG contributes negatively to $\mu_t$, prompting a reduction in OER, and vice versa.

### Conceptual Summary of GARIX

Figure C.1 provides a conceptual summary of GARIX including the dual perspectives.

**What does GARIX stand for?**

- GARIX: Global AutoRegressive Integrated time-series model with eXogenous variables and an 'equilibrium' force

**What is the aim of GARIX?** Predicting near-term changes in OER during CPB as a function of *system history*, *present state*, and *intended actions* (perspective #1):

- *System history ('past')*: Past changes in CI, Hb, $SaO_2$, temperature and OER (HTG)
- *Present state ('present')*: Current values of CI, Hb, $SaO_2$, temperature and OER (ETG)
- *Intended actions ('future')*: Planned changes in CI, Hb, $SaO_2$ and temperature (FTG)

**Alternative interpretation** - GARIX model describes OER changes as a function of *autoregressive*, *equilibrium*, and *exogenous* forces (perspective #2):

- *Equilibrium* force seeks to change oxygen consumption ($VO_2i$) - via changing OER - to ensure it matches oxygen demand ($tVO_2i$), as dictated by temperature. (ETG)
- *Autoregressive* force serves to reverse random fluctuations in OER, thus creating further stability when system is near equilibrium. (ATG)
- *Exogenous* force reflects short-term impact of changes in CI, Hb, $SaO_2$ and temperature on OER to align $VO_2i$ with $tVO_2i$. (XTG)

The GARIX model maps each of the three components in Perspectives #1 and #2 to a collection of regression terms, referred to as term groups (TGs) in the text.

**Connection between perspectives:**

- Present state of system is summarised by the equilibrium force (ETG).
- The sum of history and future term groups is equal to the sum of autoregressive and exogenous term groups, i.e., HTG + FTG = ATG + XTG.

**Model scope and idealisations (current version):**

- Model is global, i.e., parameters are shared across all patients (homogeneity assumption).
- Focus is on haemoglobin-bound oxygen (excluding dissolved oxygen).
- CI, Hb and $SaO_2$ are considered exogenous, i.e., fully controllable by the perfusionist.
- System noise is homoscedastic, i.e., random fluctuations in (logit of) OER always have the same overall strength and across all patients.
- Dependence of $tVO_2i$ on temperature follows the van't Hoff (Q10) specification: the logarithm of $tVO_2i$ is assumed to be a linear function of temperature.

Figure C.1: Conceptual summary of the GARIX model for predicting OER changes during paediatric CPB, proposed in this paper.

### D: Approximate Equivalence of van't Hoff and Arrhenius Specifications

The Arrhenius and van't Hoff formulations are the best-known parametric forms used to describe the relationship between temperature and oxygen consumption. It can be demonstrated that, if temperature changes are small compared to their absolute values, *when expressed in Kelvins*, the two specifications are approximately equivalent. In this manuscript, the van't Hoff specification has been adopted, which is consistent with the widely recognized concept of $Q_{10}$, the multiplicative increase in resting metabolic rate for every 10°C increase in body temperature.

The Arrhenius equation, after mapping the concept of chemical reaction rate to metabolism and hence oxygen consumption, would require the logarithm of oxygen consumption to be a linear function of the inverse of temperature, expressed in Kelvins ($T_k$):

$$log(VO_2 i) = a + b/T_k \tag{D.1}$$

Let's express $T_K$ in terms of its deviations ($\Delta T$) from a baseline value, $T_0$:

$$T_K = T_0 + \Delta T. \tag{D.2}$$

Equation D.1 can be rewritten as:

$$log(VO_2 i) = a + b/(T_0 + \Delta T) \tag{D.3}$$

If $\Delta T << T_0$, i.e., if changes in temperature are small compared to its baseline value, in Kelvins, then a first-order, Taylor-series expansion is used to rewrite Equation (D.3):

$$log(VO_2 i) = a + b/(T_0(1 + \Delta T/T_0)) \approx a + (b/T_0)(1 - \Delta T/T_0) = (a + b/T_0) + (-b/{T_0}^2)\,\Delta T \tag{D.4}$$

Note that the last expression on the right is a linear function of temperature change, $\Delta T$. This is simply the van't Hoff specification, i.e., logarithm of $VO_2i$ being a linear function of temperature.

### E. Model Evaluation and Diagnostics

#### Cross-Validation Procedure

##### *Repeated 5-fold Cross-Validation*

Data were split into five folds, ensuring that all observations from a single operation were contained within the same fold. The process was repeated multiple times to obtain robust estimates of model performance.

##### *Model Selection*

The history length N in the GARIX model was treated as a hyperparameter. Models with different N values were compared based on out-of-sample (OOS) R-squared performance. N=7 was selected for the final model as it provided a balance between complexity and predictive accuracy.

#### Permutation-Based Variable Importance (PVI) Analysis

##### *Objective*

To quantify the relative importance of different term groups and variables in predicting OER changes.

##### *Methodology*

Within the cross-validation framework, variables or term groups were permuted (i.e., their values shuffled) to disrupt their relationship with the outcome. The decrease in model performance due to permutation indicates the importance of the variable or term group. This method is model-agnostic and captures both linear and nonlinear relationships.

Note that random shuffling applies not to the original time-series data but to the transformed version of the data that was used in estimating the linear regression coefficients. Relatedly, when applying PVI to TGs, the same random reordering of rows was applied to all individual terms in a TG (with each term corresponding to a column in the design matrix).

#### Diagnostics

'Combined' autocorrelation functions (ACF) were computed for model residuals, and compared with Gaussian, uncorrelated noise to check for statistically significant autocorrelation. The word 'combined' refers to pooling of ACF calculations across all time-series segments in the training data. Results can be seen in the top row of Figure E.1. We see that patterns are similar; in particular, the average correlation line (red) is nearly zero in both cases, as expected. The lack of autocorrelation in the model residuals suggests that important predictors of change in $OER$ have been captured by the GARIX model.

To check for heteroscedasticity, combined ACF for the 'absolute value' of model residuals were constructed and compared to combined ACF for the absolute value of Gaussian, uncorrelated noise. Results are shown in the bottom row of Figure E.1. Here, we see a difference: While absolute residuals for the model show significantly positive mean autocorrelation for lags less than 10 minutes, no such pattern is present for the Gaussian noise. This is an indicator of heteroscedasticity, potential causes of which are explored in Discussion.

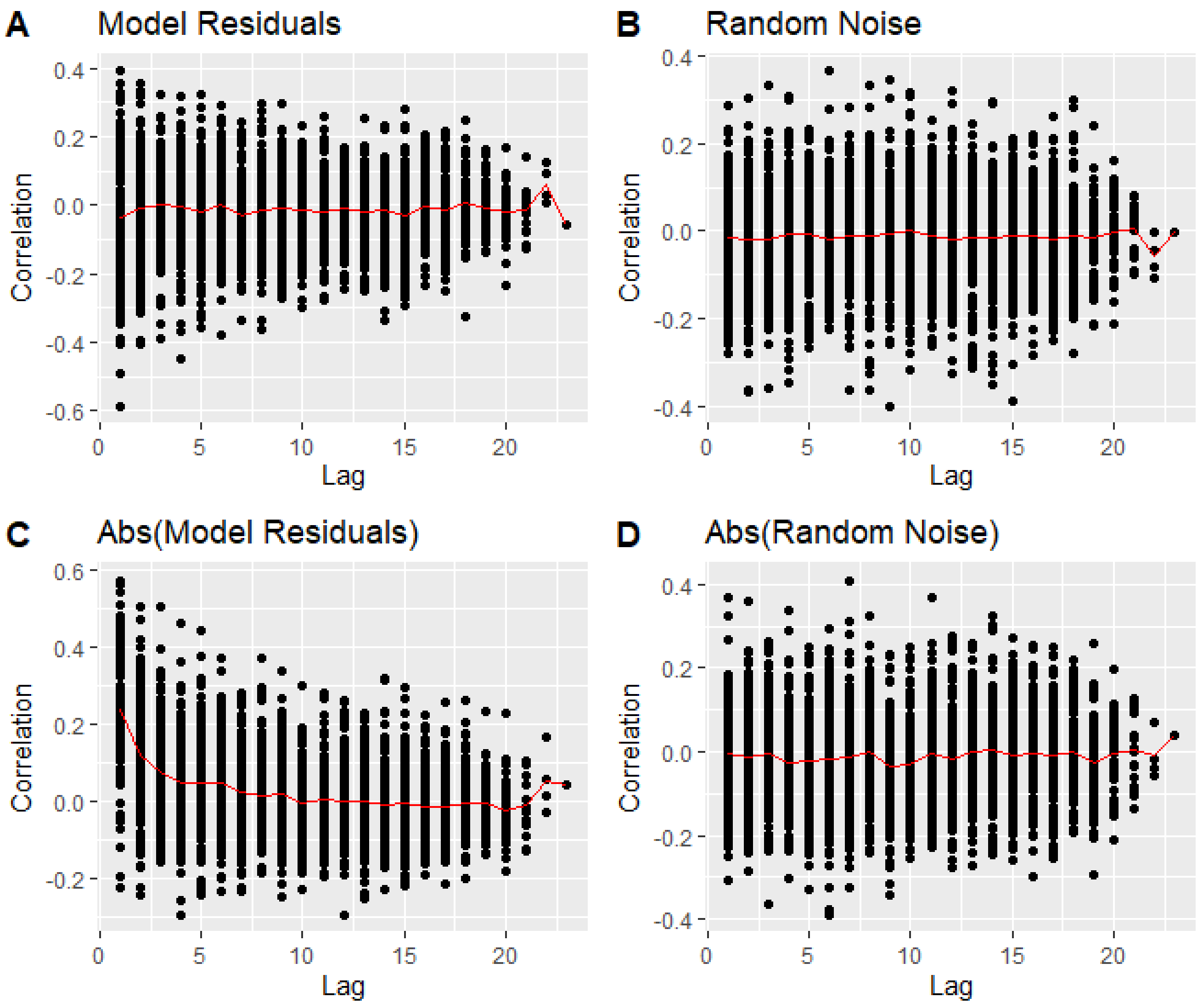


Figure E.1: Autocorrelation analysis of GARIX model residual noise. A: Combined autocorrelation plot of GARIX(7) residuals. B: Combined autocorrelation plot of random Gaussian noise for an identical time-series collection. C: Combined autocorrelation plot of the absolute values of model residuals. D: Combined autocorrelation plot of the absolute values of Gaussian noise.

### F. Statistical Significance of GARIX Coefficients

The supplementary file, *GARIX_20_model_coefficients_supp_mat.csv*, shows the list of coefficients and their statistical significance for the GARIX(20) model. Most terms have statistical significance for up to 5-10 minutes of lag, and the terms corresponding to recent history have particularly high significance. An exception is $SaO_2$, which is only highly significant at lag = 0 (corresponding to change from t to t+1).

### G. System Simulations

To understand the dynamic behaviour of the GARIX model under different scenarios, we conducted simulations using both deterministic and stochastic approaches.

#### Deterministic Simulations

**Purpose:**

- To observe the model's response to controlled changes in exogenous variables without random fluctuations.

**Procedure:**

- Initial conditions were set based on median values from the training data for CI, Hb, $SaO_2$, Temp. We then combine Equations 7-9 to solve for the steady-state OER.
- A step change was introduced in one of the exogenous variables (e.g., a 25% decrease in CI at t=0).
- The model equations were iterated forward in time, using the expected change, $\mu_t$, without adding random error.

**Outcome Measures:**

- Trajectories of OER over time.
- Contributions of each term group (ATG, XTG, ETG) to the change in OER.

#### Stochastic Simulations

**Purpose:**

- To incorporate physiological variability and assess the robustness of the model predictions under more realistic conditions.

**Procedure:**

- Similar to deterministic simulations but included the random error term $\epsilon_t$, drawn from a normal distribution with mean zero and variance $\sigma^2$ estimated from the training data.
- Multiple simulation runs were conducted to capture variability.

**Outcome Measures:**

- Visualisation of OER trajectories.
- Statistical properties such as mean absolute distance of OER from steady-state value during stochastic fluctuations.

### H. Naïve (Static) Model

As a reference point for the GARIX model, a static model for learning the dependence of tVO2i on temperature is included in the analysis. This static model assumes that $VO_2i$ is always simply equal to $tVO_2i$; in other words, both supply constraints and system dynamics are ignored. For this reason, we refer to this model as 'naïve'. As with GARIX, the relationship between $tVO_2i$ and $Temp$ is assumed to follow the van't Hoff specification:

$$log\ (VO_2i_t)\ \sim N(\beta_0 + \beta_1 T_t, \sigma^2). \tag{H.1}$$

Predictions of this model at each temperature represent the average oxygen consumption seen in the data at each temperature.

### I. Patients and Outcomes

Of a total of 963 operations, there were 854 with known AKI status. Within those, 343 had no AKI, and 286/132/93 had AKI of 1/2/3, respectively. The observed 30-day mortality rate for the entire data (0.2%) was below the expected mortality rate (1.6%), calculated according to the

PRAiS2 protocol[8], providing assurance on the reliability of data and quality of care.) Table I.1 shows data characteristics for the zero-AKI subset, which is the focus of all our subsequent analysis. Most patients (66%) were 6 months or older, and gender distribution was balanced (45% female). Most patients (67%) did not have an antenatal diagnosis.

| **Variable** | **Value** |
|---|---|
| Number of operations/patients | 343/334 |
| Median age at operation (months) | 16.9 [3.9 – 54.5] |
| Range | 0.1 – 217.4 |
| Age group (at operation) | |
| Neonates (0-30 days) | 55 (16) |
| Infants (1-6 months) | 61 (18) |
| Toddlers (6 months -3 years) | 109 (32) |
| Children (3-18 years) | 118 (34) |
| Gender | |
| Male | 187 (55) |
| Female | 156 (45) |
| Ethnicity | |
| White | 164 (48) |

| | |
|---|---|
| BAME | 135 (39) |
| <Not Available> | 44 (13) |
| Antenatal diagnosis | |
| Yes | 96 (28) |
| No | 231 (67) |
| <Not Available> | 16 (5) |

Table I.1: Summary of AKI-free patients and their outcomes. Numbers in brackets represent interquartile ranges for numeric variables, while numbers in parentheses represent percentages for discrete variables. Note that figures are reported at the operation level including for gender and ethnicity. Percentages may not add up to 100 due to rounding. BAME: Black, Asian and Minority Ethnic.
